# Modelling the public-health impact of indoor air quality interventions on respiratory virus transmission

**DOI:** 10.64898/2026.06.06.26355067

**Authors:** Adam Howes, Geetha Jeyapragasan, Richard Williamson, David Carel, Harrison Koos, Jacob L. Swett, James Montavon, Vivian Belenky, Paul Lietar, Richard Fitzjohn, Giovanni Charles, Serina Chang, Thomas Brewer, Charles Whittaker

**Author notes:** Indicates joint first-author contribution (Adam Howes, Geetha Jeyapragasan). Indicates joint senior-author contribution (Thomas Brewer, Charles Whittaker).

## Abstract

Respiratory virus transmission occurs in indoor settings where ventilation, occupancy, and dwell time determine exposure levels. Improving indoor air quality (IAQ) therefore could help reduce disease burden associated with respiratory viruses, yet its population-level impact remains poorly quantified. Here, we develop an individual-based transmission modelling framework that links within-location airborne dynamics to individual infection risk and population-level spread, whilst explicitly incorporating heterogeneity in ventilation and baseline indoor air quality across locations. We use this modelling approach to evaluate IAQ-improving interventions (air-quality interventions or AQIs), using hypothetical endemic and pandemic pathogen archetypes with properties similar to SARS-CoV-2 and influenza, and evaluate how effects on key epidemiological metrics (such as annualized incidence and epidemic final size) depend on AQI coverage, efficacy and allocation strategy. At 20% AQI intervention coverage and 80% efficacy, annualized incidence was reduced by approximately 7.2% for an endemic “SARS-CoV-2-like” respiratory virus, and 17.0% for an endemic “influenza-like” virus; at 60% coverage (80% efficacy) the reductions were 26.3% and 56.4%, respectively. Targeting AQI installation to the highest-risk locations outperformed random allocation: for SARS-CoV-2-like transmission, 20% coverage at 80% efficacy cut absolute incidence by 10.8% when targeted versus 7.2% when random; for influenza-like transmission, this comparison was 28.9% versus 17.0%. In epidemic scenarios, random installation at 40% coverage and 60% efficacy reduced final size by 23.7% (influenza-like) versus 6.3% (SARS-CoV-2-like). These results support treating clean indoor air as core public-health infrastructure and prioritising risk-based deployment of IAQ-improving interventions to maximise population-level benefit within budgetary and operational constraints.

## Background

Respiratory infections remain among the leading causes of global morbidity and mortality across the life course. Lower respiratory infections (LRIs) (predominantly pneumonia and bronchiolitis) account for over two million deaths worldwide each year, with substantial concentration of mortality in young children and older adults. In 2021 alone, LRIs were estimated to cause ∼2.18 million deaths, including ∼502,000 in children under five^1^. Viral pathogens are major contributors to this burden: seasonal influenza is responsible for around a billion infections annually, including 3–5 million severe cases and an estimated 290,000–650,000 respiratory deaths^2^; respiratory syncytial virus (RSV) drives a heavy paediatric toll, causing ∼3.6 million hospitalisations and ∼100,000 deaths in children under five each year, the vast majority in low-and middle-income countries^3^. Endemic human coronaviruses (OC43, NL63, 229E, HKU1) account for a significant proportion of experienced “common colds”, while rhinoviruses are a leading trigger of asthma and COPD exacerbations, adding substantially to healthcare use and disability^4,5^. Beyond human costs, these infections impose sizeable economic burdens through hospitalisations and productivity losses^6^. Immunity is rarely sterilising: previous work has highlighted that reinfections are common across a wide range of respiratory pathogens, with individuals experiencing seasonal coronavirus infection approximately once every 2.5 years on average^7^, influenza infection rates approaching 40% annually in some populations^8^, and SARS-CoV-2 reinfection occurring within approximately a year of prior infection^9,10^. This recurrent pattern of infection means that the cumulative burden of respiratory viral disease over the life course is substantial.

A large share of respiratory-virus transmission occurs indoors, across both households and the other settings in which individuals routinely spend a high proportion of their time and which involve sustained close proximal contact with other individuals (e.g. schools, workplaces etc.)^11^. Previous work has shown the household to be an important site of influenza virus transmission, with estimates suggesting as much as 38% of transmission might occur in households^12^. Work using COVID-19 contact tracing app data has highlighted that approximately 40% of COVID-19 transmission occurs in households, with the majority of other transmission occurring in settings recurrently visited outside the household^11^. Serological evidence from a Swiss cohort suggests around 28% of COVID-19 infections were attributable to household transmission. A corollary of these studies is that a sizeable fraction of respiratory virus transmission occurs outside the household. Indeed, evidence from population-level modelling of transmission rates across multiple European countries has demonstrated the role specifically of non-household settings in sustaining population-level spread; showing that closure of non-essential businesses and schools were associated with approximately 35% and 7% reductions in the reproduction number of COVID-19, respectively^13^. Relatedly, prior work has shown that closure of schools over holiday periods in France lead to significant reductions in influenza incidence^14^, whilst recent work examining smart thermometer data from US households has suggested as much as 70% of household viral transmission might be initiated by children infected outside of the household^15^.

Much of this transmission is thought to occur via inhalation of aerosolised virus transmitted over relatively long distances, rather than short distance transmission via direct exposure to droplets produced during symptoms such as sneezing and coughing. Previous work has demonstrated the viability of exhaled SARS-CoV-2 aerosols for transmission^16^, as well as empirical evidence from COVID-19 superspreading events during the pandemic (e.g. the Skagit Valley choir superspreading event^17^) and mathematical modelling of closed-populations on the Diamond Princess^18^ have all highlighted the significance of aerosolised SARS-CoV-2 transmission. Similar results for influenza have confirmed the presence of infectious virus in the exhaled breath of infected individuals^19^, and has suggested that as much as 50% of all transmission events may be attributable to aerosolised virus^20^.

Indoor transmission is driven, in part, by crowding and inadequate ventilation in the spaces where people spend most of their time. These conditions increase the risk of respiratory virus transmission. Although ANSI/ASHRAE Standard 62.1 sets minimum outdoor-air requirements and default occupant densities for common space types, real-world occupancy frequently exceeds design assumptions and the effective per-capita ventilation rate can drop sharply during busy periods^21,22^. Previous studies have consistently identified substandard ventilation in schools for example^23^, with measurements in recently constructed U.S. schools showing that only ∼22% of classrooms met the recommended minimum ventilation rates^24^. More broadly, previous work has highlighted that ventilation is also highly heterogeneous across setting types, and frequently insufficient. Surveys and reviews report wide distributions of air changes per hour (ACH) in homes and offices^25,26^, and that many routinely occupied rooms operate with insufficient ventilation rates^27^. Moreover, these crowding and ventilation deficits are not evenly distributed. Mobility-network analyses during COVID-19 show that disadvantaged groups tend to visit more crowded points of interest with higher modeled infection risk^28^. The structure of the built environment and how and where individuals spend their time is therefore thought to be a key driver of socio-economic inequities in disease burden. Evidence from the COVID-19 pandemic has shown that locations visited by disadvantaged racial and socioeconomic groups were, on average, more crowded and therefore associated with higher infection risk^28^, a factor in-turn likely to be driving the disparities in infection rates observed in the US during the pandemic’s earliest months^29^.

Motivated by this, a growing literature is testing practical ways to improve indoor air quality (IAQ) and, in turn, reduce respiratory-virus exposure and burden across everyday settings. Interventions aimed at achieving this goal typically span those increasing effective ventilation/air changes (such as upgrades to HVAC systems)^30^, portable particulate filtration (e.g. via HEPA filtration units)^31^, and installation of technologies aiming to directly disinfect air (such as ultraviolet germicidal irradiation approaches including far-UVC 222nm)^32^. Evidence supports the potential utility of these approaches. Lab studies have highlighted the utility of upper room UV at limiting tuberculosis transmission^33^, and that far-UVC technologies can rapidly inactivate viral bioaerosols^32^. Prior work in healthcare settings has similarly shown that portable HEPA filters with UV sterilisation can remove detectable airborne SARS-CoV-2 RNA from clinical wards^34^. In schools, recent work has shown that incorporation of air purifiers was associated with a reduction in student absences of 12.5%^35^, while other studies have highlighted associations between higher ventilation rates and decreased SARS-CoV-2 airborne transmission in school classrooms^36^ and college dormitories ^37^. Previous work has documented reduced ARI incidence in the intention-to-treat group during studies of air purifiers in residential aged care facilities^38^, and that germicidal UV light can similarly reduce ARI incidence in these settings^39^.

Despite this emerging evidence base, the majority of studies are empirical, device- and setting-specific, and frequently evaluate proximate outcomes (e.g., PM/CO₂ concentrations, pathogen RNA detection, absenteeism) within single rooms or facilities, and focus on local environments, short-horizon endpoints and specific populations. As a result, we still lack a coherent framework to quantify how indoor air interventions might alter pathogen transmission dynamics and disease burden at the population-level, and resolve their potential public-health impact under varying epidemiological conditions. Doing so will be crucial in supporting rigorous, quantitative cost–benefit analyses, as well as supporting targeting of these interventions across the diverse setting types and locations they could potentially be deployed. Motivated by this gap, we developed an individual-based epidemiological modelling framework that explicitly represents physical space and person-time in different settings (households, schools, workplaces, leisure, and community venues) and links them to within-room airborne transmission processes (ventilation, filtration, and UV inactivation). This framework integrates time-use, occupancy, and airflows with pathogen emission, dispersion, and removal. Using SARS-CoV-2 and influenza as case studies, we simulate a range of indoor-air interventions, varying efficacy, coverage, and targeting across setting types under both endemic and pandemic scenarios to estimate effects on transmission, epidemic trajectories, and health burden. Our results demonstrate substantial potential public-health benefits from improving indoor air quality, and underscore the utility of treating “clean indoor air” as core public-health infrastructure to be optimized alongside vaccines and therapeutics.

## Methods

### Population structure and data sources

We simulated a population of individuals stratified into three age groups: children, adults, and elderly **(Figure 1A)**. Household sizes **(Figure 1B)** and age composition were sampled from a 2010 San Francisco-based synthetic population produced by RTI International^40^. School enrollment sizes **(Figure 1C)** were sampled from U.S. National Center for Education Statistics 2019–2020 data^41^. Workplace sizes **(Figure 1D)** were sampled from a truncated offset power law distribution estimated previously from US-based data^42^. In the absence of detailed data and information on visit frequency and duration to other settings, we aggregated all other visits into a “leisure” category, with setting capacity **(Figure 1E)** and visit frequency **(Figure 1F)** sampled from empirically motivated negative binomial and Poisson distributions respectively.

**Figure 1:**
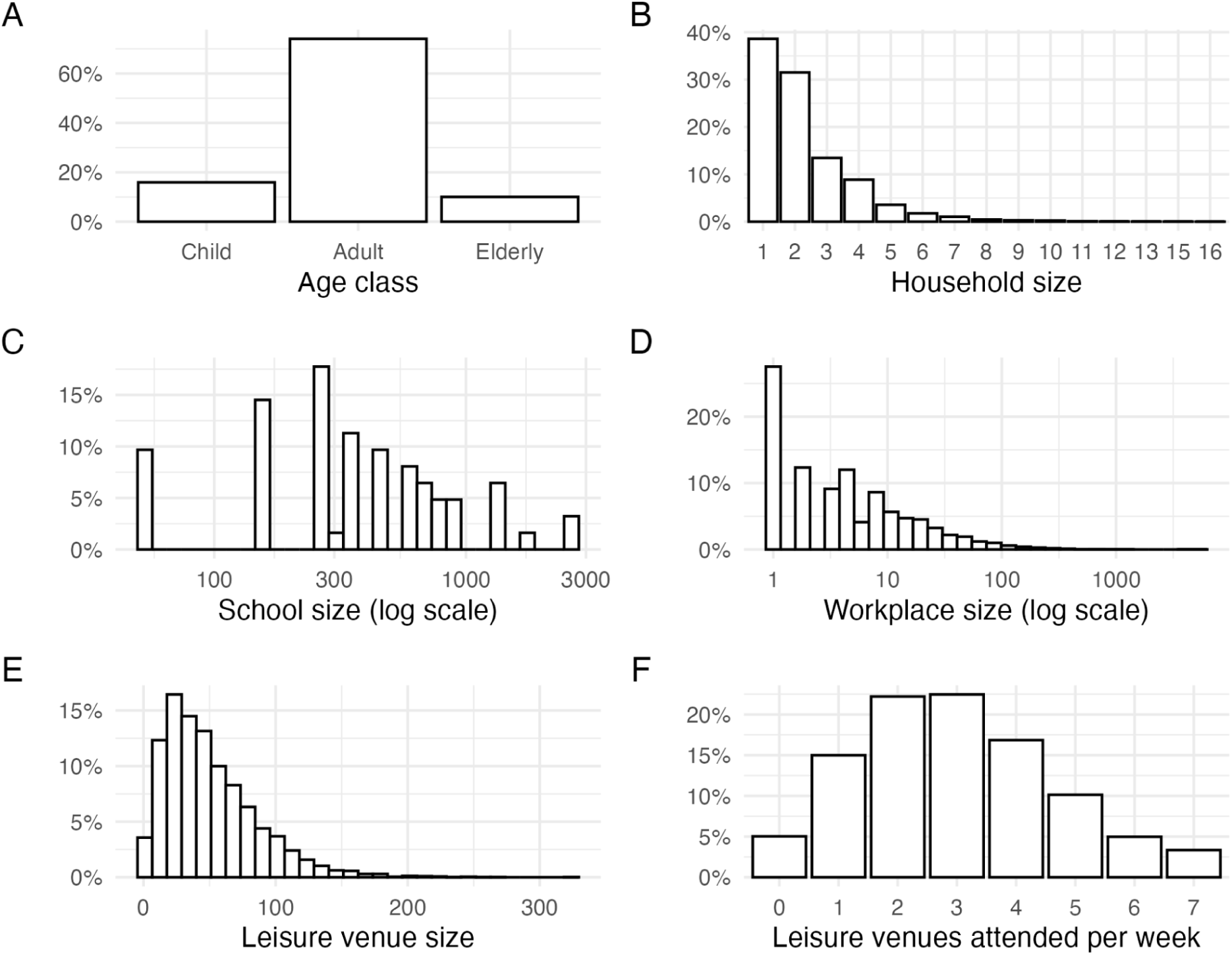
Example realised demographic and social contact setting distributions obtained for one stochastic model simulation with an illustrative population size of 200,000. **(A)** Age distribution across age classes (child, adult, and elderly). **(B)** Household size distribution showing frequency of households with 1-16 members. **(C)** School size distribution on a logarithmic scale, ranging from approximately 100 to 3,000 students. **(D)** Workplace size distribution on a logarithmic scale, ranging from 1 to over 1,000 employees. **(E)** Leisure venue size distribution showing capacity of venues attended. **(F)** Distribution of the number of leisure venues attended per week per individual.

### Overview of Individual-based modelling framework

We developed a stochastic, individual-based epidemiological model in which space is explicitly represented by assigning each individual to membership of particular locations and setting types, following the approach of Ferguson et al^42^. In what follows, we use “setting type” to denote classes of place (e.g., households, schools, workplaces, leisure venues), and “location” to index a specific instance of a given setting type (e.g., a particular household, classroom, office or leisure venue). Within the model, individuals exist in one of several disease states: susceptible, exposed (but not yet infectious), infectious, and recovered. Individuals progress through these states according to pathogen-specific natural history parameters, while acquisition of infection is governed by an exposure process that is computed based on the specific set of locations individuals visit during the course of a day. At each time step, an individual’s total force of infection (FOI) is the sum of location-specific FOIs from the locations that they visit. Individuals are assumed to visit different setting types depending on their age group.

For each location *j* belonging to setting type s, the force of infection experienced by an individual visiting that location is proportional to the fraction of visiting individuals in that location who are currently infectious. This proportion is scaled by a setting-type specific transmissibility parameter β_*s*_ and a location specific heterogeneity multiplier γ_*j*, *s*_. The former captures differences in respiratory virus transmission between setting types^11^, while the latter captures variation in occupancy, ventilation, and mixing across locations of the same setting type and is parameterised using information on ventilation rates. This construction lets us (i) control the average contribution of each setting type to transmission at the population level and (ii) represent between-location heterogeneity in transmission risk across the locations belonging to a particular setting type.

To quantify between-location “riskiness”, we use empirically documented variation in ventilation rates across locations belonging to the same setting type, and integrate these with an explicit model of within-room airborne virus concentration and a Wells–Riley dose–response model. Together, this enables us to map variation in air changes per hour, pathogen emission rates, and viral decay/inactivation into infection probabilities as a function of dwell time. This provides a consistent means to link variation in baseline ventilation rates to location-specific transmission risk, and encode ventilation/filtration/UV parameters as multiplicative reductions in location-specific force of infection. This framework (described in further detail below) is implemented in R as an open-source package^43^, built using the *individual* R package; package documentation, vignettes, and source code are publicly available. Further model details are provided below, but see https://mrc-ide.github.io/helios/ for full details.

### Transmission dynamics and calculation of individual force of infection

Following the approach of Ferguson et al^42^, the force of infection (FOI) experienced by an individual *i* at time *t* is decomposed across the location *j* and setting type *s* as follows

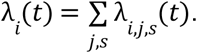

With the location and setting type-specific FOI calculated as

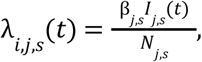

where *I*_*j*,*s*_(*t*) is the number of infectious individuals in location *j* belonging to setting type *s*, *N*_*j*,*s*_ is the occupancy in that particular location, and β_*j*,*s*_ = γ_*j*,*s*_β_*s*_ is a transmissibility parameter. The term β_*s*_ represents a setting-type specific transmissibility, parameterised in order to recapitulate empirically observed patterns of the proportion of transmission for influenza and SARS-CoV-2 occurring across different setting types, while γ_*j*,*s*_ ∼ *Lognormal*(0, σ_*s*_) captures between-location heterogeneity in transmission risk arising from variation in ventilation rates and occupancy density, for a particular setting type (e.g. schools). We set between-setting transmission rate ratios based on estimates from prior work about how overall transmission is apportioned between settings^11,44^

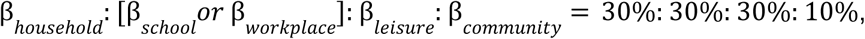

with β_*school*_ *or* β_workplace_ depending on the age-class an individual belongs to, and with the sum of β_*s*_ values scaled to achieve overall levels of transmission consistent with reproduction number estimates for the specific pathogen archetypes modeled (see below). Households, workplaces, schools and leisure settings are explicitly represented within the modelling framework; “community” transmission is used to represent residual infection risk occurring in locations or contexts outside these settings.

### Indoor air quality motivated variation in riskiness between locations

To represent variation in transmission riskiness across locations *j* belonging to the same setting type *s*, γ_*j*,*s*_ , we followed the approach of Blatchley et al^45^, modeling airborne pathogen concentration *C*(*t*) using an ordinary differential equation

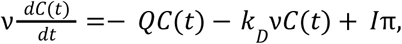

where ν is room volume, *Q* is air flow rate, *k*_*D*_ = 0.64*h*^−1^ is a pathogen decay constant^45^, and π = 27 virions per person per hour is the emission rate^45^. We assumed room volume ν = *ND*_*s*_*H* where *N* is the maximum room occupancy, *D*_*s*_ is floor area (*m*^2^) per person (*D_household_* = 20, *D_school_* = 3. 33, *D_workplace_* = 10, *D_leisure_* = 2^46^), and *H* = 2. 5*m* is the room height. For a given infectious viral concentration *C*, the probability of an individual becoming infected conditional on some time *t* spent in that location is calculated based on the Wells-Riley equation^47,48^, as

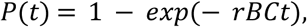

with breathing rate *B* = 0.45*m*^3^_h_^−1^ and infection probability per virion *r* =1.37×10^−2 45,49^. We calculated upper and lower bounds of infection probability using setting-specific ranges of air changes per hour derived from a review of setting type and location ventilation rates^27^, and assuming individuals spend 12 hours of the day in households, 8 hours in schools/workplaces, 2.5 hours at leisure venues, and 1.5 hours in community settings. The distribution of location-specific variation in infection risk derived from the variation in ventilation rates was then used to parameterise a lognormal distribution of setting type and location specific riskiness values, with median 1 and with log-standard deviation σ_*log, s*_.

### Modelling the impact of air quality interventions

We modeled the impact of generic air quality interventions (AQIs) that either modify the physical number of air changes per hour (ACH) or increase the rate at which exhaled virus becomes inactive (thereby decreasing the total virus concentration in a room, in what is typically referred to as the “effective air changes per hour” or eACH). Given substantial heterogeneity between different AQIs in terms of changes to eACH and dependence on the specific ventilation dynamics of each space, as well as the initial, exploratory nature of this work, we made the simplifying assumption that modeled AQIs installed in a given setting would reduce the location-specific FOI multiplicatively by efficacy *E*_*j*,*s*_ and modify the FOI as follows

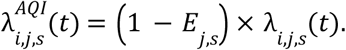

With this formulation, we then carried out a suite of sensitivity analyses varying AQI efficacy, coverage level (i.e. the target proportion of floor area receiving AQI installation), and allocation strategy (randomly distributed across locations vs. targeted to highest-risk eligible locations). In all instances, AQIs were assumed to be installed in schools, workplaces and leisure facilities only, with the assumption that private residences (i.e. households) would not have these installed.

### Disease and AQI scenario modelling

We simulated two pathogen archetypes possessing the epidemiological properties of influenza and SARS-CoV-2, including the natural history (average incubation period of 2 days and an average duration of infectiousness of 4 days for SARS-CoV-2; average incubation period of 1 day, average duration of infectiousness of 2 days for influenza), and reproduction number (approximately *R*_0_ ≈ 1. 5 for influenza and *R*_0_ ≈ 2. 5 for SARS-CoV-2). For both pathogen archetypes, we carried out a suite of simulations assuming both endemic transmission (i.e. in which transmission is recurring and individuals are routinely reinfected due to declining immunity level, and which matches the dynamics of SARS-CoV-2 and influenza currently); and an alternative set of scenarios (“epidemic” scenarios) in which a novel pathogen with properties similar to existing influenza and SARS-CoV-2 viruses emerges into the population. We simulated both endemic and epidemic scenarios for each pathogen archetype under multiple AQI strategies: no AQI (baseline), random allocation of AQI to locations, and targeted allocation to the highest-risk eligible locations. Each scenario was replicated 25 times to account for stochastic variation between model runs. For endemic scenarios, we computed annualized disease incidence and average prevalence of active infection over the time-period simulated. Simulations were run for 20 years with an assumed mean duration of immunity of 365 days, and interventions were introduced following the end of the 10th year, across indoor settings including workplaces, schools, and leisure venues. The period spanning years 5–7 served as the baseline, and years 15–17 as the post-intervention phase for purposes of evaluation. For epidemic scenarios, we examined epidemic final size, the time to the peak of the epidemic, and peak infection incidence as metrics to assess AQI impact.

## Results

### AQI installation can achieve substantial reductions in endemic respiratory virus infection incidence burden, though impact depends on coverage and efficacy

Across the explored scenarios, AQI installation reduced annualized disease incidence and the prevalence of active infections across both pathogen archetypes considered, though overall impact depended strongly on coverage (defined as the proportion of non-household floor area receiving AQI installation) and the efficacy of the AQI (defined as the proportion reduction in the force of infection experienced by a susceptible individual in the space where the AQI was installed). Assuming random allocation of AQIs to locations, at 20% coverage and 80% efficacy, annualized disease incidence decreased by 7.2% (range 5.0%-9.2% across 25 stochastic simulations) for the SARS-CoV-2-like pathogen and by 17.0% (range 10.0%-25.5%) for the influenza-like pathogen **(Figure 2A & 2D)**. Active infection prevalence declined by 6.8%, from 1.5% to 1.4% on average for SARS-CoV-2 and declined by 18.8%, from 0.6% to 0.5% for influenza on average **(Figure 2B & 2E)**. Increasing coverage or efficacy both led to significantly increased reductions in infection incidence **(Figure 2C & 2F)**. For example, keeping efficacy fixed at 80% and increasing coverage to 60%, reductions in annualized incidence were 26.3% (range 23.4%-30.0%) and 56.4% (range 49.1%-64.5%) for SARS-CoV-2 and influenza, respectively. Similarly, fixing coverage at 20%, average reductions in annualized incidence were 3.1%, 4.8% and 7.2% for the SARS-CoV-2-like pathogen at 40%, 60% and 80% assumed efficacy, respectively, and 8.8%, 11.6% and 17.0% for the influenza-like pathogen.

**Figure 2:**
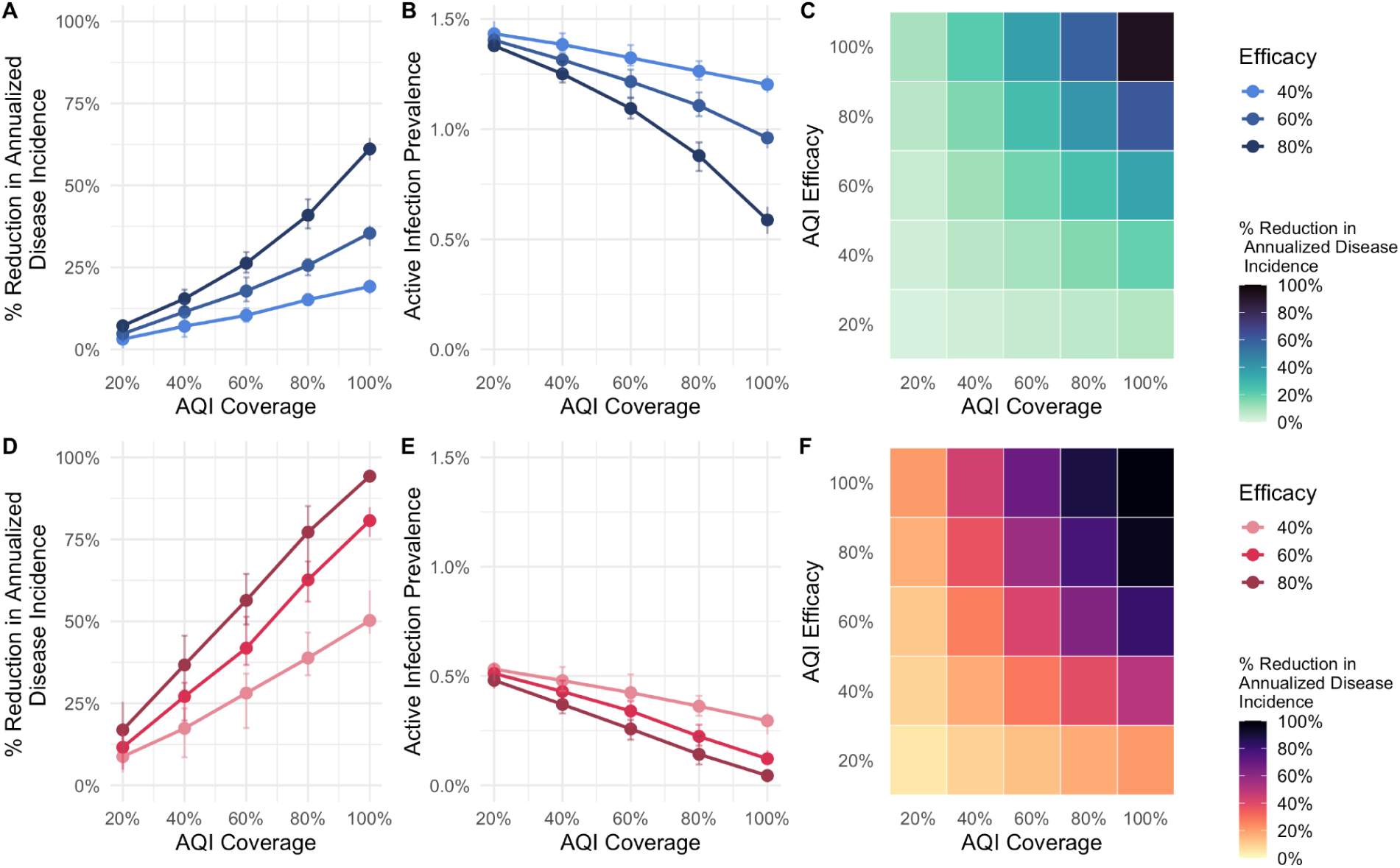
Impact of AQI installation implementation on infection incidence. **(A)** The percentage reduction in annual incidence of infection (y-axis) and how this varies with AQI coverage (x-axis), for the SARS-CoV-2-like archetype. Coloured lines represent different assumptions about the efficacy of installed AQIs at reducing onward transmission. Error bars indicate the range of impact observed across 25 stochastic simulations. **(B)** Active infection prevalence (y-axis) and its dependence on AQI coverage for the SARS-CoV-2-like archetype. (**C)** Heatmap showing percentage reduction in annualized disease incidence across AQI coverage and efficacy. **(D)** As for **(A)**, but for the influenza-like archetype. **(E)** As for **(B)**, but for the influenza-like archetype. **(F)** As for **(C)**, but for the influenza-like archetype.

### AQI installation has a larger impact on “influenza-like” pathogens than on “SARS-CoV-2-like” pathogens

Across all intervention scenarios explored, AQI installation produced greater proportional reductions in disease for the influenza-like archetype than for the SARS-CoV-2-like archetype **(Figure 2)**. Across equivalent coverage and efficacy levels ranging from 40% to 80%, reductions in annualized disease incidence for influenza were 1.9–2.8× higher than those observed for SARS-CoV-2. Similarly, reductions in active infection prevalence were approximately 2.0–2.3× higher for influenza than for SARS-CoV-2 across the same range of coverage and efficacy. These differences reflect the higher sensitivity of pathogens with lower R₀ and shorter generation times to reductions in per-contact transmission risk, suggesting that AQI installation would have particularly strong effects on influenza-like endemic pathogens.

### Specifically targeting high-risk settings substantially increases the impact of AQI installation

Across all efficacy and coverage combinations explored, targeting installation of AQI to the highest-risk settings consistently produced significantly greater reductions in infection incidence compared to scenarios where installation was done randomly (i.e. installed in settings irrespective of transmission risk) **(Figure 3A)**. For example, at 20% coverage and 80% efficacy, targeting installation to highest-risk locations reduced annual incidence by an average of 10.8% (range 9.2%-13.2%) for the SARS-CoV-2 archetype compared with 7.2% under random installation (range 5.0%-9.2%), a 50% relative increase in impact **(Figure 3B)**. Similar patterns were observed for the influenza archetype, with targeted deployment achieving a 28.9% (range 21.8%-34.0%) reduction compared to the random installation achieving a 17.0% at equivalent coverage and efficacy, a 71.0% relative increase in impact **(Figure 3C)**. Targeting AQI installation to the riskiest settings enables the same reduction in annualized incidence to be achieved with lower levels of overall coverage. For example, for the SARS-CoV-2 archetype, a 30% reduction in annualized incidence required slightly above 60% AQI coverage under random implementation, while targeted coverage achieved the same reduction at only ∼45% coverage **(Fig. 3B)**. This disparity was even more pronounced for the influenza archetype – at 80% efficacy, a 30% reduction in annualized incidence was reached with only 20% targeted coverage, compared with 40% coverage when AQI installation was done at random. A 50% reduction in infection incidence required ∼40% targeted coverage versus ∼60% random coverage **(Figure 3E)**. These results indicate that targeting high-risk settings can reduce the coverage required for equivalent population-level impact and substantially improve the efficiency of AQI deployment.

**Figure 3:**
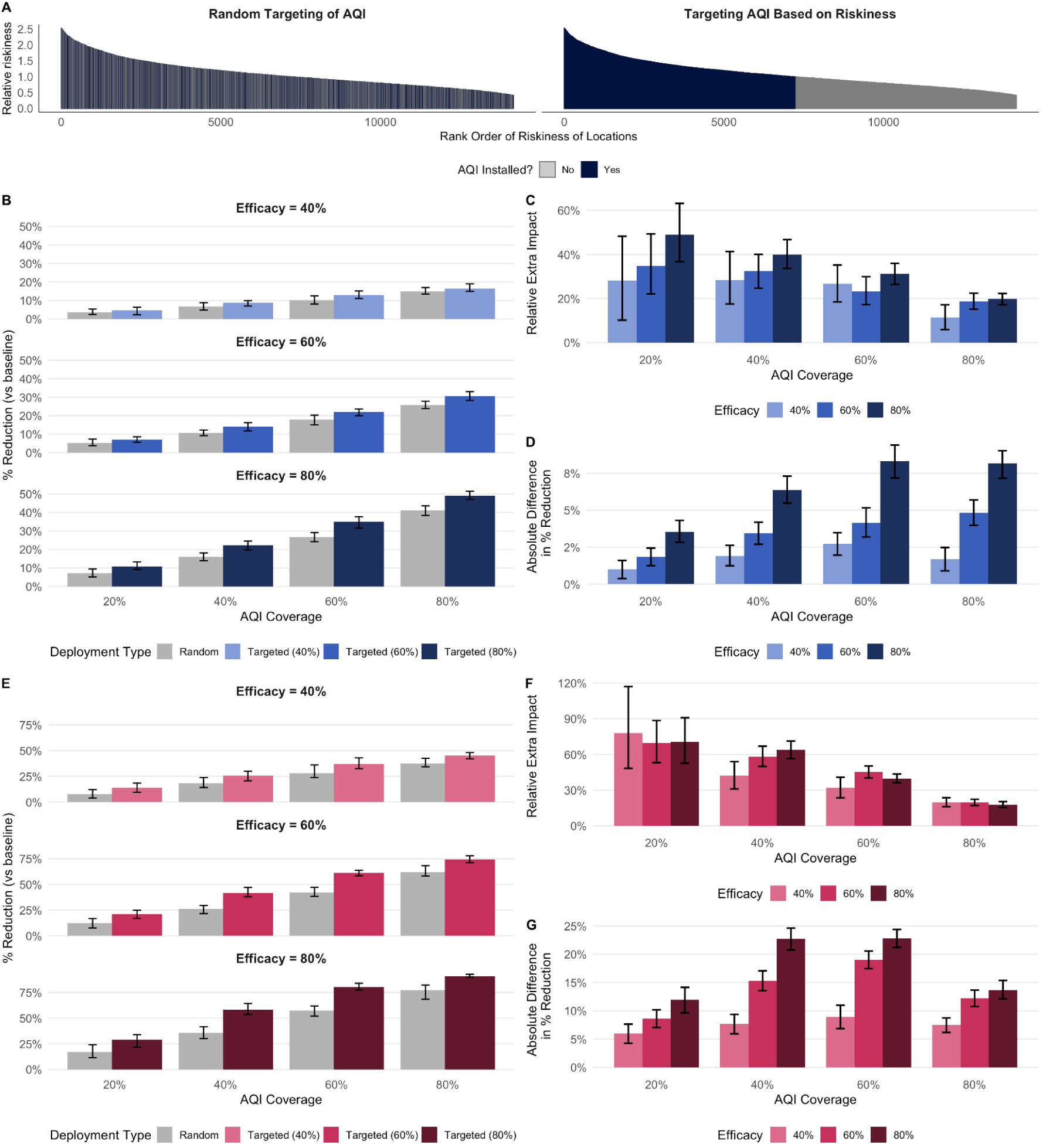
Comparison of the impact achieved under random and risk-based AQI installation and deployment strategies. **(A)** Schematic contrasting random selection of locations with risk-based targeting when 50% of locations are equipped; grey bars indicate locations without AQIs, dark blue bars indicate locations with AQIs installed. **(B)** Percentage reduction in annual incidence (y-axis) across increasing coverage (x-axis) for random (grey) versus targeted (blue) implementation, under alternative assumptions about AQI efficacy. Bars show the mean; error bars show the range across 25 stochastic simulations. **(C)** Relative added impact of targeted versus random deployment. **(D)** Absolute difference in percentage reduction between targeted and random strategies. **(E)** As for (B) but for influenza-like archetype. **(F)** As for (C) but for influenza-like archetype. **(G)** As for (D) but for influenza-like archetype.

### Risk-based targeting provided the greatest relative benefit at low coverage levels

Across both pathogen archetypes, the relative benefit of targeting AQI installation based on riskiness (compared to random allocation) was greatest at low coverage levels and declined as coverage increased (and thus the two strategies converge; being identical when 100% of locations have AQI installed) **(Figure 3C & 3F)**. At 20% coverage and 60% efficacy, targeting increased the relative population-level impact of AQI installation by 22.1% - 49.3% relative to random deployment for the SARS-CoV-2 archetype, and by 53.1% - 88.6% for the influenza archetype. The benefit remained substantial at moderate coverage but diminished at higher levels, with relative extra impacts falling to 19.7% (range 17.0%-22.4%) for SARS-CoV-2 and 18.8% (range 15.2%-22.5%) for influenza at 80% coverage. This is due to the fact that as coverage increases, there is a greater overlap between locations selected by random and riskiness-based targeting. The absolute difference in percentage reduction between random and targeted allocation strategies followed a similar pattern **(Figure 3D & 3G)** peaking at mid-range coverage before converging at high coverages (as in these scenarios, most locations received AQIs). These results indicate that targeting high-risk settings provides the largest proportional advantage when deployment resources are limited.

### AQI installation could also significantly impact pathogen transmission dynamics and infection incidence during future pandemic scenarios

Next, we evaluated the impact of AQIs on infection incidence across two archetypal epidemic scenarios, again approximating influenza and SARS-CoV-2 as archetypes. Across both epidemic archetypes, higher efficacy or wider coverage led to reductions in infection incidence across all metrics considered. Without intervention, the peak incidence of the influenza archetype was 18.6 per 1,000 (range 15.2-23.2) with a final epidemic size of 61.5% of total population (range 57.6%-66.1%), whereas the SARS-CoV-2 archetype yielded a peak incidence of 40.4 per 1,000 (range 37.1-44.6) and a final epidemic size of 89.7% of total population (range 88.7%-91.2%). As with our results examining endemic scenarios, the impact of AQI installation was more pronounced in the influenza archetype. Random AQI installation at 40% coverage and 60% efficacy reduced the final epidemic size by 23.7% (range 18.9%-31.8%), compared with only 6.3% (range 4.5%-8.2%) for the SARS-CoV-2 archetype under the same settings **(Figure 4A & 4E)**.

**Figure 4:**
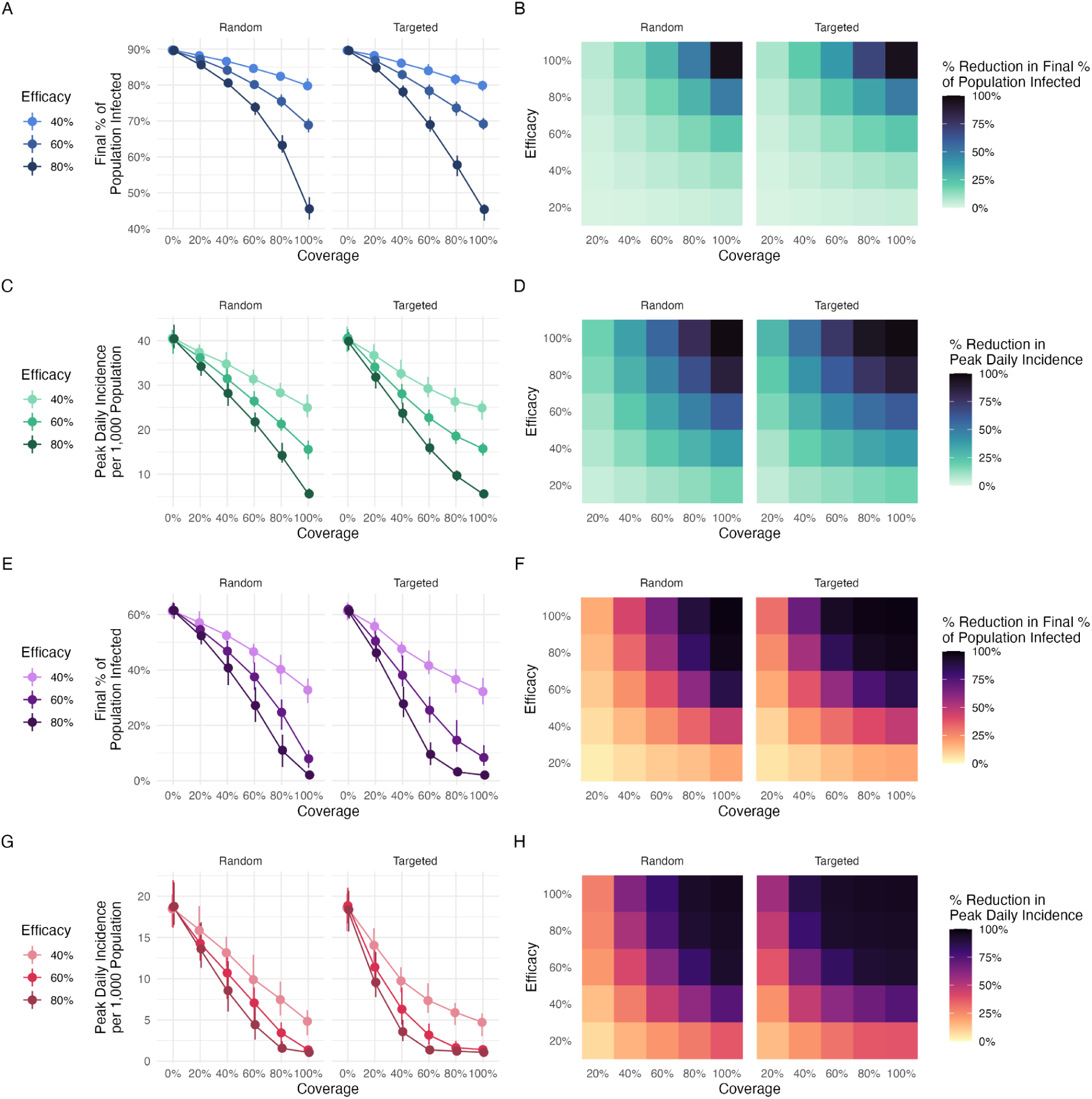
Analyses of the impact of air quality interventions on infection incidence across epidemic scenarios. **(A)** Final epidemic size in terms of percentage of the population infected for the SARS-CoV-2 archetype, with point-ranges showing the mean and simulated range across coverage levels for random versus targeted deployment; line and point colors denote intervention efficacy. **(B)** Heatmap of final epidemic size for SARS-CoV-2, providing sensitivity analysis across a wider range of values. **(C)** As for **(A)**, but for the peak incidence measured in terms of daily infections per 1,000 population. **(D)** As for **(B)**, but for the peak incidence. **(E)** Final epidemic size for the influenza archetype. **(F)** Heatmap of final epidemic size for the influenza archetype. **(G)** As for **(E)**, but for the peak incidence. **(H)** As for **(F)**, but for the peak incidence.

Increasing either efficacy or coverage yielded similar increases in the impact on epidemic peak, timing and final size. For the influenza archetype, expanding randomly allocated coverage from 40% to 60% at 40% efficacy lowered the final epidemic size by 11.1%, approximately matching the 10.7% from instead increasing efficacy from 40% to 60% while holding coverage at 40% **(Figure 4E)**.

As with our endemic scenario results, targeted AQI strategies achieved higher reductions in infection incidence at lower coverage levels than the random strategy. For the influenza archetype at 60% efficacy, the random strategy required 60% coverage to reach an average peak daily incidence of 7.06 per 1,000, while the targeted strategy achieved lower results with only 40% coverage, producing an average peak daily incidence of 6.30 per 1,000 **(Figure 4G)**. This pattern also held at lower coverage levels: the random strategy’s result at 40% coverage (average peak daily incidence of 10.7 per 1,000) was comparable to the targeted strategy’s outcome at just 20% coverage, where peak daily incidence was 11.4 per 1,000 **(Figure 4G)**. In these cases, taking a targeted approach allowed a 20% reduction in the coverage required to achieve a given infection incidence reduction.

## Discussion

Our results highlight the significant impact that widespread improvements to indoor air quality (IAQ) could potentially have on respiratory-virus transmission and associated disease burden. Across the pathogen archetypes explored here, we highlight the potential for widespread introduction of AQIs to yield significant reductions in endemic disease burden as coverage and efficacy increase, with larger gains when interventions are targeted to higher-risk locations rather than distributed at random. In epidemic scenarios, AQIs reduce peak incidence and population-level spread. Our modeled results suggest that AQIs of modest efficacy and population coverage are unlikely to control pandemics driven by pathogens with moderate to high *R*_0_ (e.g. above 1.5) on their own. However, widespread presence of these technologies would i) slow epidemic progression and spread, buying valuable time for countermeasure development, healthcare capacity surging, etc.; and ii) reduce the need for community-based mitigation measures (such as school or business closures) to control spread, which are typically highly costly from a socio-economic perspective. These conclusions are consistent with current practice guidance that positions ventilation, filtration, and ultraviolet germicidal irradiation as foundational layers of protection, and with policy frameworks that formalize minimum performance targets for infectious-aerosol control^50^. Taken together, our analyses support interventions improving indoor air as capable of supporting burden reduction while underscoring that realized impact will hinge on coverage, targeting, maintenance, and other factors relating to their practical deployment^51^.

In endemic settings, we find that reductions in infection incidence scale with both coverage and efficacy of AQIs, with larger absolute and relative effects for the influenza-like archetype modeled than for the SARS-CoV-2-like archetype. Near the epidemic threshold of *R* = 1, standard epidemic theory implies strongly non-linear behaviour, with small proportional reductions in transmission translating into disproportionately large changes in equilibrium incidence and prevalence when the pathogen reproduction number is closer to this threshold^52,53^. Baseline reproduction numbers for seasonal and pandemic influenza typically lie closer to the epidemic threshold of *R* = 1 (typically between 1.2 and 1.4 for seasonal influenza^54^), whereas early SARS-CoV-2 estimates were higher (often *R* > 2. 5)^55^; a difference which we captured in the way we parameterised the pathogen archetypes presented here. For example, coverage of 40% of non-household floor area with AQIs with efficacy 60% led to 26.2% and 10.2% reductions in annual infection incidence for influenza and SARS-CoV-2 respectively. Crucially, because overall annual infection incidence for our SARS-CoV-2-like pathogen is higher, this still translates into sizeable reductions in absolute burden, even though it remains a (relatively) smaller fraction of total burden. In both cases, achieving substantial impact (e.g. >30% reduction) was contingent on comparatively high levels of coverage and/or efficacy: to achieve 30% reduction at assumed 60% efficacy required ∼∼90% and ∼45% coverage for the SARS-CoV-2-like and influenza-like pathogen, respectively.

In our modeled epidemic scenarios, installed AQIs reduced the final epidemic size and peak incidence, as well as lengthening the time to peak, with more pronounced effects again for the influenza-like archetype than for SARS-CoV-2-like scenarios. Our modeled results show that AQIs of modest efficacy and population coverage are unlikely to achieve containment of the most highly transmissible pathogens on their own. However, these results also indicate that these measures could still play a role in both reducing and slowing epidemic growth and associated population-level spread, buying time for the development of countermeasures under accelerated (but still delayed relative to epidemic onset) timelines, as with the Coalition for Epidemic Preparedness’ 100 Days Mission. Moreover, by lowering baseline transmission in public and community venues, IAQ improvements may also reduce reliance on disruptive measures such as prolonged school or business closures, which carry documented and persistent educational and macroeconomic costs^56–58^. Together, these results position clean indoor air as a supportive component of layered epidemic control, even if not a standalone panacea for it.

Despite these promising results, an important limitation of the analyses presented here is that sizeable and reliable burden disease reductions arise most clearly at high coverage (typically >30% of targeted buildings). Widespread and comprehensive improvement of indoor air quality across such a large stock of buildings is likely to be very expensive, and whether such levels can be achieved (especially in the short-term) remains an important practical question that we do not address here. Important to note however, is that the costs of different AQI technologies remain deeply uncertain (and likely highly variable), and no rigorous cost-effectiveness analysis has yet compared AQI interventions to alternative disease control measures (such as population-level vaccination). Without such analyses, whether AQI investments compare favourably to the per-person costs of other population-level control measures (such as vaccinating a comparable population fraction) remains an open question. Irrespective, maximizing AQI impact for a given coverage level or budget is likely to remain crucial. Our results indicate that targeting IAQ interventions to settings that contribute disproportionately to onward transmission yields larger population-level benefits than random allocation at the same coverage, with the advantage of targeting greatest when coverage is low and diminishing as coverage approaches saturation. This pattern is consistent with previous theoretical work highlighting that strategic allocation of general transmission reducing measures outperforms random allocation and that the two converge only at very high coverage^59,60^, as well as empirical evidence that a minority of venues account for a large share of infections^28^ and that transmission is highly overdispersed^61,62^.

Targeted allocation of IAQ interventions to the riskiest settings would enable significantly more impact to be achieved even at lower levels of coverage; but consistently and comprehensively identifying these highest-risk settings remains challenging. The exact location in which infection occurs is not typically observed and difficult to identify via traditional contact-tracing based methods, especially in the context of asymptomatic or pre-symptomatic transmission (as for COVID-19^63,64^) and constraints on the resourcing of contact tracing infrastructure^65^. Additionally, transmission risk is strongly context dependent: beyond ventilation (which we use as a proxy for risk in the analyses presented here and which represents a significant limitation of our work), factors such as occupancy, duration of co-presence, vocalization and activity intensity shape emission and exposure^66,67^. Resolving patterns of transmission and risk at finer spatial granularity is therefore crucial. Previous work has highlighted the promise of digital contact tracing as a way to provide time-stamped proximity records that strengthen case–context linkage when recall is incomplete and exposure windows span multiple venues^68^. Similarly, previous work has highlighted that aggregated mobile network data can quantify venue-level person-time and crowding from anonymized device movements to estimate setting-specific exposure and risk^28^. More broadly, scalable sensing systems for proxies of poor ventilation and thus high transmission risk (such as CO₂) could also comprehensively characterize ventilation across locations and identification of persistently under-ventilated spaces^69,70^.

Indeed, a major limitation of this work is the simplified representation of the physical spaces that individuals visit and the contact structures they form within them. Although we differentiate setting types and locations and parameterize their relative contributions using estimates derived from previous work^42^, we do not explicitly model within-room spatial mixing, airflow patterns, or the dynamic co-location networks that arise from realistic daily schedules and repeated encounters. As a result, our framework cannot capture the fine-grained paths by which people move between venues during a day, the distribution of dwell times at specific points of interest, or the concurrency of contacts that shapes exposure risk. Future extensions that integrate time-use data to constrain schedules and dwell times such as the American Time User Survey^71^ or mobility network data from mobile phones or similar would enable a more realistic reconstruction of non-household transmission opportunities.

We also do not explicitly differentiate shorter-range, droplet-mediated transmission (more closely determined by the specific interpersonal contacts made within a location or particular room) from longer-range aerosol inhalation, which depends more on room airflows and ventilation^66^. Instead, we represent AQI effects via fixed efficacies that implicitly combine both the intrinsic performance of the intervention and the proportion of total transmission that is avertible by air-quality modification. The comparative contributions of these different pathways currently remains poorly resolved, and evidence suggests that relative contribution may vary across pathogens and contexts. Infectious influenza virus has been detected in fine exhaled aerosols from symptomatic cases^19^ and infectious SARS-CoV-2 aerosols have been shown to be emitted by COVID-19 patients^72^. For RSV however, aerosol spread has been demonstrated experimentally but remains less well characterized than droplet and contact routes^73^. Better resolving these distinct mechanisms and integrating them into the same framework would be a valuable direction for future work.

A related limitation concerns our representation of within-room transmission dynamics. Although our framework advances prior individual-based models by explicitly linking setting-level ventilation and riskiness to exposure via a Wells-Riley-based mechanism^74^, we assume a steady-state, well-mixed approximation for each location that fails to capture fine-scale within-location granularity in airflow dynamics and spread of exhaled infectious virus throughout the space^75^. We also neglect inter-room airflows and building-scale transport, representing between-location variation through stochastic heterogeneity in overall transmission risk rather than explicitly coupling that altered risk to changes or differences in airflow. These simplifications were chosen for computational tractability at population scale, but they omit mechanisms that can materially influence risk in specific spaces^76,77^. Further refining our current representation of indoor space and adding more realistic spatial structure and airflow within a particular location, such as computational fluid dynamics (CFD) models with our existing IBM would be valuable.

Another limitation is that we do not represent specific AQI technologies or undertake health-economic evaluations, limiting our ability to assess the cost-effectiveness of interventions aiming to improve IAQ. This choice reflects the present evidence base: randomized and quasi-experimental studies of venue-level AQIs remain limited in number, heterogeneous in design, and mixed in outcomes. For example, trials in aged-care facilities showed no clear reduction in acute respiratory infections with in-room HEPA filtration and only modest, aggregate effects with germicidal UV in long-term care^38,39^, while reviews underscore sparse transmission-endpoint data and variable implementation quality^51^. Formal cost-effectiveness evidence is similarly limited: some analyses during the COVID-19 period suggest circumstances where portable filtration or ventilation upgrades could be cost-effective, but these findings depend on assumptions about device performance, usage patterns, energy costs, and background incidence that vary widely across settings^78,79^. Rigorous evaluation of comparative effectiveness remains a priority, but is methodologically challenging: trial design is complicated by the need for large sample sizes, because individuals typically spend only a fraction of their infectious period in any single trial venue, so venue-specific interventions act on only a limited share of exposure; this dilutes detectable effects and increases required sample sizes; a feature that is further complicated by the possibility of averted infections occurring via other routes, in other locations individuals visit where IAQ interventions are not installed. Generating such evidence is therefore crucial. Integrating it into population-level models of spread and epidemiological impact will facilitate rigorous cost-effectiveness analyses across payer perspectives (societal, health-system, building owner or operator, employer or school district, etc.), decision contexts (e.g. time horizon, discounting choices etc.), and co-benefits (such as reduced PM_2.5_ exposure and productivity or attendance gains).

Despite these limitations, our work highlights the substantial burden reduction that could be achieved by improving indoor air quality at scale, across both endemic and epidemic contexts. Realizing this benefit will depend on implementation quality and coverage across diverse building stocks, sustained operation and maintenance, and alignment with performance-based standards that specify equivalent clean air delivery and infection-risk management modes. It will also require practical guidance and workforce capacity to assess and upgrade ventilation, filtration, and air disinfection in priority settings. Further research should quantify real-world effectiveness for specific technologies in varied venues, generate comparative- and cost-effectiveness evidence across payer perspectives, and develop measurement systems that resolve venue-level risk in near-real time.

**Table 1:** Pathogen archetype natural history parameters.

| Parameter | SARS-CoV-2-like | Influenza-like | Source(s) |
| --- | --- | --- | --- |
| Approximate basic reproduction number ($R_0$) | 2.5 | 1.5 | Liu et al. 2020; Biggerstaff et al. 2014 |
| Mean incubation period (days) | 2 | 1 | Model assumption |
| Mean infectious period (days) | 4 | 2 | Model assumption |
| Mean duration of immunity (days) | 365 | 365 | Model assumption |

**Table 2:** Model parameters.

| Parameter | Baseline value | Range | Source |
| --- | --- | --- | --- |
| Population structure |  |  |  |
| Population size | 50,000 | Fixed | Model assumption |
| Age groups | 3 (child, adult, elderly) | Fixed | Model assumption |
| Household size distribution | ~1-16 members | Sampled: synthetic population | RTI International 2010 (Rineer et al. 2025) |
| School enrollment distribution | ~100-3,000 students | Sampled: empirical data | US NCES 2019-2020 |
| Workplace size distribution | ~1-1,000+ employees | Sampled: truncated offset power-law distribution | Ferguson et al. 2005 |
| Leisure venue size distribution | ~1-300 | Sampled: negative binomial distributed | Model assumption |
| Transmission dynamics |  |  |  |
| Household transmission proportion | 30% | Fixed | Ferretti et al. 2024; Ferguson et al. 2006 |
| School/Workplace transmission proportion | 30% | Fixed | Ferretti et al. 2024 |
| Leisure transmission proportion | 30% | Fixed | Ferretti et al. 2024 |
| Community transmission proportion | 10% | Fixed | Model assumption (residual transmission) |
| Between-location heterogeneity | Median 1 with setting-specific standard deviation | Sampled: lognormal distribution | Corsi et al. |
| Indoor air quality and ventilation |  |  |  |
| Pathogen decay constant ($k_d, h^{-1}$) | 0.64 | Fixed | Blatchley & Cui 2023 |
| Viral emissions per person per hour | 27 | Fixed | Blatchley & Cui 2023 |
| Breathing rate ($m^3 h^{-1}$) | 0.45 | Fixed | Blatchley & Cui 2023 |
| Infection probability per virion | 0.0137 | Fixed | Blatchley & Cui 2023; Killingley et al. 2022 |
| Room height ($m$) | 2.5 | Fixed | Model assumption |
| Household occupancy density (per $100m^2$) | 5 | Fixed | ASHRAE 62.1 (Kelechava 2022) |
| School occupancy density (per 100m<sup>2</sup>) | 30 | Fixed | ASHRAE 62.1 (Kelechava 2022) |
| Workplace occupancy density (per 100m<sup>2</sup>) | 10 | Fixed | ASHRAE 62.1 (Kelechava 2022) |
| Leisure occupancy density (per 100m<sup>2</sup>) | 50 | Fixed | ASHRAE 62.1 (Kelechava 2022) |
| Air changes per hour (ACH) | Setting-specific | Sampled: Lognormal per setting | Corsi et al. |
| Air quality interventions |  |  |  |
| Settings with AQI | Schools, workplaces, leisure | Fixed | Model assumption |
| Settings without AQI | Households, community | Fixed | Model assumption |

**Table 3:** Simulation parameter settings.

| Setting | Values |
| --- | --- |
| Pathogen archetype | SARS-CoV-2-like; Influenza-like |
| AQI efficacy | 20%, 40%, 60%, 80%, 100% |
| AQI coverage | 0%, 20%, 40%, 60%, 80%, 100% |
| AQI deployment | Random; Risk-targeted |
| Transmission regime | Endemic; epidemic |
| Stochastic replicates | 25 |

## Data Availability

Code to reproduce our results are available from https://github.com/petal-code/helios-paper

