## Supplementary Information for "Modelling the public-health impact of indoor air quality interventions on respiratory virus transmission"

**4 Computational information** **11**

### 1 Introduction

We developed an individual-based susceptible-exposed-infectious-recovered (SEIR) transmission model to simulate the effects of interventions that modify indoor air quality on the transmission of an airborne pathogen. The model incorporates both age structure and social mixing patterns across multiple locations within different types of setting. To assess the impact of interventions that modify air quality, we altered the force of infection across locations. The model is available as an open-source R package called `helios` (`https://github.com/mrc-ide/helios/`), which was developed using the `individual` R package (Charles and Wu, 2021).

### 2 Model structure

#### 2.1 Population

We simulated a human population consisting of a fixed number of individuals, $i = 1, \ldots, N$. The population was divided into three age groups: children, adults, and the elderly (Section 2.2). Each individual was assumed to visit different settings throughout the course of a day (Section 2.3). We did not explicitly represent the amount of time that individuals spend in each setting. Instead, we used empirical data for the proportion of transmission that occurs by setting (Section 2.5).

#### 2.2 Age

There are three age groups individuals can be assigned to: children, adults and the elderly. Individuals remain in their assigned age group for the full simulation. The age group an individual is assigned to informs the settings they are assigned to. For example, children are assigned to schools and adults are primarily assigned to workplaces.

We assigned individuals to age groups using data from a synthetic household panel representative of San Francisco, California. These data were developed in 2010 by RTI International to inform disease modelling (Wheaton and RTI, 2014). We sampled households from this synthetic panel with replacement. If the final household sampled caused the population size to exceed $N$, the excess individuals were dropped from the model.

#### 2.3 Spatial structure

To capture social mixing patterns, we assumed that individuals divide their time between the following types of setting: household, workplace, school, leisure, and community.

##### 2.3.1 Households

Each individual was assigned to a household which remained fixed for the duration of the simulation. Household assignments were generated jointly with age assignments as described in Section 2.2.

##### 2.3.2 Community

We assumed a background rate of mixing between all individuals intended to represent an overall community setting. A background force of infection term applies to all individuals regardless of the location.

##### 2.3.3 Schools

All children were assumed to attend school. School assignments were fixed throughout the duration of the simulation. To inform the distribution of school sizes, we used national-level US school data for 2019-2020 from the National Center for Education Statistics, available from de Brey et al. (2023). We sampled with replacement from this list of school sizes. All schools categorised as prekindergarten, elementary, middle, or secondary and high were included. School sizes were provided in ranges (e.g. "500 to 599"), so we used the midpoint (e.g. 550). We modified the final sample such that the sum of the total school sizes equalled the population size of children.

Some adults were also assumed to attend schools as staff, with each school having a fixed staff-to-student ratio of 1:20. Adults assigned as staff in a school were not assigned a workplace location (Section 2.3.4).

###### 2.3.4 Workplaces

All adults (who are not school staff) are assumed to attend workplaces. The number of employees in each workplace was sampled from a truncated offset power statistical distribution with cumulative distribution function

$$\mathbb{P}(n > m) = \frac{\left(\frac{1+N/a}{1+m/a}\right)^c - 1}{(1+N/a)^c - 1}, \quad \text{for } m \leq N. \tag{1}$$

This equation was based on that of Ferguson et al. (2006), which in turn is based on data from the United States (U.S. Bureau of Labor Statistics, 2003). We used the parameters $a = 5.36$ and $c = 1.34$, and truncated draws at 10% of the total adult population. We modified the final sample such that the sum of all workplace sizes equalled the eligible workforce size. Adults were randomly assigned to each of these workplaces and assumed to visit the same workplace each day.

###### 2.3.5 Leisure venues

All individuals are eligible to attend leisure venues. The maximum occupancy sizes of the leisure venues were sampled from a truncated negative binomial distribution. The negative binomial parameters were mean 50 and overdispersion 2, with all samples greater than 10% of the total population truncated. We modified the final sample such that the sum of all leisure venue capacities equalled the population size.

Individuals were assigned to visit different leisure venues each day (or sometimes not to visit any leisure venues on a given day). The number of leisure venues visited by each individual per week was sampled from a Poisson distribution with rate 3. The samples were truncated at seven, corresponding to a daily leisure visit. Particular leisure venues visited each day were drawn without replacement from the list of all leisure venues, with probability proportional to venue capacity.

##### 2.4 Disease states

We used an individual-based implementation of an SEIR compartmental model with susceptible (S), exposed (E), infectious (I), and recovered (R) disease states. Discrete timesteps with a step size of $\Delta t = 0.5$ days were used in the model. At each timestep $t$, individuals transition between states stochastically. A susceptible individual transitions to exposed with probability $1 - \exp(-\lambda_i(t)\,\Delta t)$, exposed individuals become infectious with probability $1 - \exp(-\frac{1}{\sigma}\,\Delta t)$, and infectious individuals recover with probability $1 - \exp(-\frac{\Delta t}{\delta})$. In the endemic model, recovered individuals return to the susceptible state with probability $1 - \exp(-\frac{\Delta t}{\omega})$. The term $\lambda_i(t)$ is the total force of infection experienced by an individual $i$ at time $t$. The terms $\sigma$, $\delta$, and $\omega$ are the mean residence times in the exposed, infectious, and recovered states respectively.

#### 2.5 Force of infection

The total force of infection $\lambda_i(t)$ is the sum of the location-specific force of infections terms that individual $i$ experiences across all settings they visit in that timestep

$$\lambda_i(t) = \sum_{j,s} \lambda_{i,j,s}(t). \tag{2}$$

The force of infection an individual experiences at a given location was modelled as

$$\lambda_{i,j,s}(t) = \frac{\beta_{j,s} I_{j,s}(t)}{N_{j,s}}, \tag{3}$$

where $\beta_{j,s}$ is a transmissibility parameter for location $j$ in setting $s$ that integrates consideration of both the amount of time spent in a location and the inherent "riskiness" of a location, $I_{j,s}(t)$ is the number of individuals who are infected in location $j$ of setting $s$ at time $t$, and $N_{j,s}$ is the total number of individuals in location $j$ of setting $s$. Transmissibility varied between locations in a setting according to

$$\beta_{j,s} = \gamma_{j,s} \beta_s,$$

where $\beta_s$ is the average riskiness of the setting (Section 2.5.1) and $\gamma_{j,s}$ are location-specific relative riskiness parameters (Section 2.5.2).

##### 2.5.1 Variation in risk between settings

We set $\beta_s$ using a 3:3:3:1 ratio of setting-specific transmissibility parameters $\beta_{\text{household}}$ : [$\beta_{\text{school}}$ or $\beta_{\text{workplace}}$] : $\beta_{\text{leisure}}$ : $\beta_{\text{community}}$. These parameters are based on data on the proportion of transmission that occurs within each setting (Ferguson et al., 2006; Bi et al., 2021; Ferretti et al., 2024). We set the sum over $\beta_s$ based on the particular pathogen in order to achieve a given effective reproduction number (Section 2.7).

| Setting | $\sigma_{log,s}$ | Riskiness ratio $R_s$ |
|---|---|---|
| School | 0.3544 | 4.75 |
| Workplace | 0.5072 | 6.35 |
| Household | 0.0871 | 2.5 |
| Leisure | 0.4278 | 5.5 |

Table 1: Setting-specific riskiness distribution parameters

##### 2.5.2 Variation in risk within settings

To capture heterogeneity in transmission risk between locations in a particular setting type, each location $j$ of setting type $s$ was assigned a riskiness multiplier $\gamma_{j,s}$ at the start of each simulation. The multipliers were drawn from a truncated lognormal distribution

$$\gamma_{j,s} \sim \text{TruncLognormal}\left(\mu_{\text{log}} = 0,\ \sigma_{\text{log},s},\ \text{min} = \tfrac{1}{\sqrt{R_s}},\ \text{max} = \sqrt{R_s}\right).$$

Each multiplier scales the FOI for all individuals at that location relative to the setting-level transmissibility $\beta_s$. A log-mean of 0 centers the distribution at one, such that $\beta_s$ represents the transmission rate at an average location. The distribution was scaled using setting-specific log-standard deviation parameters $\sigma_{\text{log},s}$ and truncated based on setting-specific riskiness ratios $R_s$. We informed these parameters (Table 1) empirically using a Wells-Riley model of airborne transmission (Blatchley III and Cui, 2023).

In particular, the steady-state concentration of infective virus in an enclosed space is

$$C_{ss} = \frac{I\pi}{\alpha\nu}, \qquad \alpha = \frac{Q}{\nu} + k_D,$$

where $\nu = N D_s H$ is room volume ($N$ maximum occupants; floor area per person $D_s$; room height $H = 2.5$m), $Q$ is volumetric airflow ($Q/\nu$ gives ACH), $k_D = 0.64\,\text{h}^{-1}$ is the viral decay rate, $I = 1$ is the number of infectors, and $\pi = 27\,\mathrm{h}^{-1}$ is the infectious-unit emission rate (Blatchley III and Cui, 2023). Floor area per person was parameterised as: $D_{\text{household}} = 20$, $D_{\text{school}} = 3.33$, $D_{\text{workplace}} = 10$, $D_{\text{leisure}} = 2$, taken from American Society of Heating, Refrigerating and Air-Conditioning Engineers (2022) (Table 6.1) for non-household settings and assumed for households. The probability of infection after dwell time $t$ is

$$P_i(t) = 1 - \exp(-rBC_{ss}t),$$

where $B = 0.45\mathrm{m}^3$ per hour is the inhalation rate of virus particles and $r = 1.37 \times 10^{-2}$ is the probability of infection per infective-virion inhaled (Killingley et al., 2022; Blatchley III and Cui, 2023). We evaluated $P_i$ at the minimum and maximum ACH for each setting type (Corsi, 2006), using dwell times of 12 h (households), 8 h (schools and workplaces), and 2.5 h (leisure). The resulting riskiness ratios $P_i(\mathrm{ACH}_{\min})/P_i(\mathrm{ACH}_{\max})$ ranged from 2.5-fold (households) to 6.35-fold (workplaces). For each setting and associated set of riskiness ratios, we estimated the value of $\sigma_{log,s}$ that best defined a truncated log-normal with bounds $[\frac{1}{\sqrt{R_s}}, \sqrt{R_s}]$. For each setting and riskiness ratio, we set $\sigma_{log,s}$ to the value that places approximately 80% of the probability mass of the truncated lognormal distribution within the range $[\frac{1}{\sqrt{R_s/2}}, \sqrt{R_s/2}]$, with 10% of the mass in each tail, identified through numerical optimization.

#### 2.6 Intervention model

We modelled the impact of air quality interventions (AQIs) as a multiplicative reduction to the force of infection (Equation 3) such that

$$\lambda^{\text{AQI}}_{i,j,s}(t) = (1 - E_s) \times \lambda_{i,j,s}(t),$$

where $E_s$ is applied uniformly to all locations within setting $s$ that receive an AQI. We considered efficacy values of 20%, 40%, 60%, 80%, and 100%. We applied AQIs across schools, workplaces and leisure settings, excluding households as they are unlikely to have AQIs installed.

Coverage was defined as the target proportion non-household floor area receiving AQI installation. Two allocation strategies were considered: random and targeted. Under random allocation, locations were selected uniformly at random from all non-household settings until the cumulative covered floor area met or exceeded the coverage target. Under targeted allocation, locations were ranked by their riskiness $\gamma_{j,s}$, with AQIs installed in locations of descending order of riskiness until the cumulative covered floor area met or exceeded the coverage target.

#### 2.7 Pathogen archetypes

We simulated pathogen archetypes with transmission intensities approximating those of influenza and SARS-CoV-2. To parameterise pathogens that approximate these archetypes, we used the `finalsize` package (Gupte et al., 2024) to determine the final epidemic size, as a proportion of the total human population, expected for a pathogen with an $R_0$ equivalent to influenza ($R_0 \approx 1.5$) or SARS-CoV-2 ($R_0 \approx 2.5$). We then determined the setting-specific transmissibility parameters ($\beta_s$) required to generate an epidemic with a final size equivalent to each of the pathogen archetypes, maintaining the 3:3:3:1 ratio of setting-specific transmissibility parameters (Table 2).

| Parameter | Influenza | SARS-CoV-2 |
|---|---|---|
| mean exposed duration | 1 | 2 |
| mean infectious duration | 2 | 4 |
| $\beta_{\text{household}}$ | 0.207 | 0.24 |
| $\beta_{\text{school}}$ | 0.207 | 0.24 |
| $\beta_{\text{workplace}}$ | 0.207 | 0.24 |
| $\beta_{\text{leisure}}$ | 0.207 | 0.24 |
| $\beta_{\text{community}}$ | 0.069 | 0.08 |

Table 2: Pathogen parameters required to generate epidemic final sizes for pathogens with $R_0$ values equivalent to those of influenza and SARS-CoV-2.

### 3 Model outputs

To quantify the impact of AQIs on disease transmission, we summarised each model run using a set of epidemiological metrics.

#### 3.1 Endemic scenarios

For endemic scenarios, we simulated 20 years of transmission dynamics. The first four years served as a burn-in, with years 5-7 used as the baseline period without interventions. AQIs were implemented at the beginning of year 11, with outcomes assessed during years 15-17. We considered two output metrics: the annualized disease incidence and active infection prevalence.

Annual incidence is the proportion of the population newly infected over one year,

$$\text{Annual incidence} = \frac{1}{N} \sum_{t \in \mathcal{T}} E_{\text{new}}(t), \tag{4}$$

where $E_{\text{new}}(t)$ is the number of new exposures at timestep $t$, $N$ is the total population, and

$\mathcal{T}$ denotes the set of timesteps within the year.

Active infection prevalence is the proportion of the population that is currently exposed or infectious. We summarise this by its time-average over the year,

$$\text{Mean active infection prevalence} = \frac{1}{|\mathcal{T}|} \sum_{t \in \mathcal{T}} \frac{E_t + I_t}{N}, \tag{5}$$

where $E_t$ and $I_t$ are the numbers of exposed and infectious individuals at timestep $t$, and $|\mathcal{T}|$ is the number of timesteps in the year.

##### 3.2 Epidemic scenarios

For epidemic scenarios, we computed the final epidemic size and the peak incidence.

The final epidemic size is the proportion of the population infected over the course of the outbreak,

$$\text{Final epidemic size} = \frac{R_{\text{final}}}{N}, \tag{6}$$

where $R_{\text{final}}$ is the total number of recovered individuals at the end of the epidemic.

Peak incidence is the maximum per-capita rate of new exposures observed in any single timestep,

$$\text{Peak incidence} = \frac{1}{N} \max_{t \in \mathcal{T}} E_{\text{new}}(t). \tag{7}$$

### 4 Computational information

Simulations were performed on Imperial College London's Department for Infectious Disease Epidemiology (DIDE) high-performance computing cluster using the `hipercow` package (FitzJohn et al., 2025).
